# Racial and ethnic differences in COVID-19 vaccine hesitancy and uptake

**DOI:** 10.1101/2021.02.25.21252402

**Authors:** Long H. Nguyen, Amit D. Joshi, David A. Drew, Jordi Merino, Wenjie Ma, Chun-Han Lo, Sohee Kwon, Kai Wang, Mark S. Graham, Lorenzo Polidori, Cristina Menni, Carole H. Sudre, Adjoa Anyane-Yeboa, Christina M. Astley, Erica T. Warner, Christina Y. Hu, Somesh Selvachandran, Richard Davies, Denis Nash, Paul W. Franks, Jonathan Wolf, Sebastien Ourselin, Claire J. Steves, Tim D. Spector, Andrew T. Chan, on behalf of the COPE Consortium

## Abstract

**Background:** Racial and ethnic minorities have been disproportionately impacted by COVID-19. In the initial phase of population-based vaccination in the United States (U.S.) and United Kingdom (U.K.), vaccine hesitancy and limited access may result in disparities in uptake.

**Methods:** We performed a cohort study among U.S. and U.K. participants in the smartphone-based COVID Symptom Study (March 24, 2020-February 16, 2021). We used logistic regression to estimate odds ratios (ORs) of COVID-19 vaccine hesitancy (unsure/not willing) and receipt.

**Results:** In the U.S. (*n*=87,388), compared to White non-Hispanic participants, the multivariable ORs of vaccine hesitancy were 3.15 (95% CI: 2.86 to 3.47) for Black participants, 1.42 (1.28 to 1.58) for Hispanic participants, 1.34 (1.18 to 1.52) for Asian participants, and 2.02 (1.70 to 2.39) for participants reporting more than one race/other. In the U.K. (*n*=1,254,294), racial and ethnic minorities had similarly elevated hesitancy: compared to White participants, their corresponding ORs were 2.84 (95% CI: 2.69 to 2.99) for Black participants, 1.66 (1.57 to 1.76) for South Asian participants, 1.84 (1.70 to 1.98) for Middle East/East Asian participants, and 1.48 (1.39 to 1.57) for participants reporting more than one race/other. Among U.S. participants, the OR of vaccine receipt was 0.71 (0.64 to 0.79) for Black participants, a disparity that persisted among individuals who specifically endorsed a willingness to obtain a vaccine. In contrast, disparities in uptake were not observed in the U.K.

**Conclusions:** COVID-19 vaccine hesitancy was greater among racial and ethnic minorities, and Black participants living in the U.S. were less likely to receive a vaccine than White participants. Lower uptake among Black participants in the U.S. during the initial vaccine rollout is attributable to both hesitancy and disparities in access.

## Introduction

Severe acute respiratory syndrome coronavirus 2 (SARS-CoV-2) and the COVID-19 pandemic have claimed 2.4 million lives among nearly 110 million confirmed cases worldwide.^1^ The speed and urgency with which multiple vaccines have been authorized for use in the United States (U.S.),^2, 3^ the United Kingdom (U.K.),^4–6^ and elsewhere^7^ represent an unrivaled scientific achievement. However, there is a critical need for effective vaccine delivery to realize the promise of ending the pandemic. Logistical hurdles and supply chain difficulties have plagued the early phase of a massive global vaccination campaign, particularly in the U.S.^8^ As of February 17, 2021, only 17 doses of vaccine per 100 individuals have been administered in the U.S. compared with 24 per 100 in the U.K.^9^

Racial and ethnic minorities are at particularly increased risk of COVID-19, its related complications, and death.^10–12^ Nonetheless, eligibility for most vaccine programs have prioritized health care workers (HCW), older adults and those with comorbidities, but have not considered race or ethnicity.^13^ In addition to concerns over fairness and availability, a substantial barrier to uptake in minority communities is vaccine hesitancy, rooted in ongoing discrimination and prior injustices that have resulted in deeply seated mistrust of the medical system.^14, 15^

The U.S. and U.K. have diverse populations that have been comparably stricken by the COVID-19 pandemic. In contrast to the U.K., which has centralized vaccine delivery through the National Health Service, U.S. efforts have been led by fragmented state and local health authorities that have not routinely collected information on race, ethnicity, or hesitancy.^16^ In both countries, there have been reports of racial/ethnic disparities in vaccine uptake, but specific data across a broad community-based sample, particularly in the U.S., are lacking. To assess the real-world impact of the initial phase of these vaccination programs, we used an established smartphone-based data collection tool^17^ to conduct a comparative population-based cohort study to examine country-specific variation in racial and ethnic disparities in vaccine willingness and receipt.

## Methods

### Study design and participants

We performed a cohort study in the U.S. and U.K. using the COVID Symptom Study (CSS) smartphone application developed by Zoe Global Ltd. in collaboration with researchers at the Massachusetts General Hospital, King’s College London, Lund University, and Uppsala University.^17^ This research study was approved by the Mass General Brigham Human Research Committee (Institutional Review Board Protocol 2020P000909) and King’s College London Ethics Committee (REMAS ID 18210).

Beginning December 10, 2020–two days after the first authorized vaccine administration to a member of the U.K. public,^19^ we introduced a questionnaire to U.K. participants assessing whether they received a vaccine dose. Starting January 7, 2021 in the U.S. and U.K. (study baseline), we collected information on participant willingness to obtain a COVID-19 vaccine (yes/no/unsure) if it was offered to them and any suspected vaccine-related symptoms. For those unsure about or unwilling to receive a vaccine, we queried their underlying reasons (**Table S1**).

**Table 1.** Baseline characteristics of study participants by race and ethnicity according to country of enrollment

|  | United States |  |  |  |  |
| --- | --- | --- | --- | --- | --- |
|  | White<br>(n=76286) | Black<br>(n=2468) | Hispanic<br>(n=3838) | Asian<br>(n=3602) | Other/More than<br>one<br>(n=1194) |
| **Age (years)** | 60.9 (15.0) | 58.5 (16.0) | 50.5 (18.1) | 55.8 (20.3) | 56.9 (17.1) |
| <25 | 1804 (2.4) | 110 (4.5) | 341 (8.9) | 370 (10.3) | 58 (4.9) |
| 25-34 | 4035 (5.3) | 149 (6.0) | 563 (14.7) | 376 (10.4) | 106 (8.9) |
| 35-44 | 6603 (8.7) | 210 (8.5) | 597 (15.6) | 409 (11.4) | 136 (11.4) |
| 45-54 | 8716 (11.4) | 341 (13.8) | 646 (16.8) | 445 (12.4) | 163 (13.7) |
| 55-64 | 15970 (20.9) | 635 (25.7) | 664 (17.3) | 434 (12.0) | 237 (19.8) |
| ≥65 | 39158 (51.3) | 1023 (41.5) | 1027 (26.8) | 1568 (43.5) | 494 (41.4) |
| **Female sex** | 50523 (66.2) | 1825 (73.9) | 2421 (63.1) | 2193 (60.9) | 773 (64.7) |
| **BMI (kg/m<sup>2</sup>)** | 27.0 (6.0) | 30.0 (7.0) | 27.9 (6.4) | 25.1 (5.1) | 28. (6.7) |
| <18.5 | 31208 (40.9) | 582 (23.6) | 1337 (34.8) | 1900 (52.7) | 404 (33.8) |
| 18.5-24.9 | 1480 (1.9) | 32 (1.3) | 91 (2.4) | 139 (3.9) | 18 (1.5) |
| 25-29.9 | 25206 (33.0) | 815 (33.0) | 1261 (32.9) | 1085 (30.1) | 419 (35.1) |
| ≥30 | 18392 (24.1) | 1039 (42.1) | 1149 (29.9) | 478 (13.3) | 353 (29.6) |
| **Comorbidities** |  |  |  |  |  |
| Diabetes | 3770 (4.9) | 274 (11.1) | 215 (5.6) | 272 (7.6) | 74 (6.2) |
| Heart Disease | 5988 (7.8) | 165 (6.7) | 219 (5.7) | 263 (7.3) | 102 (8.5) |
| Lung Disease | 2261 (3.0) | 76 (3.1) | 59 (1.5) | 68 (1.9) | 46 (3.9) |
| Kidney Disease | 1392 (1.8) | 63 (2.6) | 65 (1.7) | 79 (2.2) | 37 (3.1) |

|  | United States |  |  |  |  |
| --- | --- | --- | --- | --- | --- |
|  | White<br>(n=76286) | Black<br>(n=2468) | Hispanic<br>(n=3838) | Asian<br>(n=3602) | Other/More than<br>one<br>(n=1194) |
| Cancer | 2202 (2.9) | 55 (2.2) | 80 (2.1) | 79 (2.2) | 27 (2.3) |
| Education | 45.5 (18.4) | 36.5 (17.5) | 38.9 (18.9) | 47.0 (17.7) | 43.1 (19.1) |
| Income | 81106 (31527) | 68086 (28329) | 74883 (30427) | 89684 (31559) | 78042 (30526) |
| Current/prior smoking | 22254 (29.2) | 723 (29.3) | 992 (25.8) | 838 (23.3) | 392 (32.8) |
| Healthcare worker | 5985 (7.8) | 226 (9.2) | 346 (9.0) | 273 (7.6) | 105 (8.8) |
| Prior COVID-19 | 4542 (6.0) | 204 (8.3) | 514 (13.4) | 192 (5.3) | 89 (7.5) |

|  | United Kingdom |  |  |  |  |
| --- | --- | --- | --- | --- | --- |
|  | White<br>(n=1204721) | Black<br>(n=9615) | South Asian<br>(n=17628) | Middle East/East<br>Asian<br>(n=7689) | More than<br>one/other<br>(n=14641) |
| **Age (years)** | 55.0 (15.4) | 47.4 (15.6) | 49.6 (15.1) | 49.3 (15.4) | 46.7 (15.7) |
| <25 | 46399 (3.9) | 920 (9.6) | 816 (4.6) | 369 (4.8) | 1462 (10.0) |
| 25-34 | 92413 (7.7) | 1312 (13.6) | 2021 (11.5) | 1055 (13.7) | 2022 (13.8) |
| 35-44 | 159125 (13.2) | 1676 (17.4) | 4063 (23.0) | 1710 (22.2) | 3122 (21.3) |
| 45-54 | 238482 (19.8) | 2186 (22.7) | 4391 (24.9) | 1668 (21.7) | 3172 (21.7) |
| 55-64 | 306131 (25.4) | 2323 (24.2) | 3145 (17.8) | 1404 (18.3) | 2811 (19.2) |
| ≥65 | 362171 (30.1) | 1198 (12.5) | 3192 (18.1) | 1483 (19.3) | 2052 (14.0) |
| **Female sex** | 711693 (59.1) | 5642 (58.7) | 9883 (56.1) | 4438 (57.7) | 9016 (61.6) |
| **BMI (kg/m<sup>2</sup>)** | 26.6 (6.0) | 27.7 (7.2) | 25.5 (5.7) | 25.3 (6.0) | 26.2 (6.7) |
| <18.5 | 517674 (43.0) | 3477 (36.2) | 8779 (49.8) | 4119 (53.6) | 7033 (48.0) |
| 18.5-24.9 | 31567 (2.6) | 376 (3.9) | 796 (4.5) | 334 (4.3) | 590 (4.0) |
| 25-29.9 | 404507 (33.6) | 3087 (32.1) | 5422 (30.8) | 2131 (27.7) | 4265 (29.1) |
| ≥30 | 250973 (20.8) | 2675 (27.8) | 2631 (14.9) | 1105 (14.4) | 2753 (18.8) |
| **Comorbidities** |  |  |  |  |  |
| Diabetes | 39168 (3.3) | 429 (4.5) | 1233 (7.0) | 275 (3.6) | 461 (3.1) |
| Heart Disease | 48828 (4.1) | 258 (2.7) | 832 (4.7) | 300 (3.9) | 440 (3.0) |
| Lung Disease | 25971 (2.2) | 155 (1.6) | 294 (1.7) | 118 (1.5) | 226 (1.5) |
| Kidney Disease | 11089 (0.9) | 112 (1.2) | 206 (1.2) | 69 (0.9) | 128 (0.9) |
| Cancer | 17046 (2.1) | 121 (1.8) | 151 (1.3) | 71 (1.4) | 129 (1.4) |
|  | White<br>(n=1204721) | Black<br>(n=9615) | South Asian<br>(n=17628) | Middle East/East<br>Asian<br>(n=7689) | More than<br>one/other<br>(n=14641) |
| **Education** | 7.1 (2.5) | 6.5 (2.6) | 7.24 (2.50) | 7.5 (2.4) | 7.4 (2.5) |
| **Income** | 7.0 (2.5) | 5.7 (2.7) | 6.41 (2.65) | 6.6 (2.6) | 6.5 (2.6) |
| **Current/prior smoking** | 272021 (22.6) | 2210 (23.0) | 2711 (15.4) | 1533 (19.9) | 3110 (21.2) |
| **Healthcare worker** | 47300 (3.9) | 636 (6.6) | 1365 (7.7) | 404 (5.3) | 611 (4.2) |
| **Prior COVID-19** | 72555 (6.0) | 1020 (10.6) | 1762 (10.0) | 625 (8.1) | 1242 (8.5) |

Census-level data on education used the education score and income score for the Index of Multiple Deprivation, respectively. N (percentages) presented for categorical variables. Values are means (SD) for continuous variables. Values of polytomous variables may not sum to 100% due to rounding
Abbreviations: BMI (body mass index), m (meter), kg (kilogram)

### Ascertainment of racial/ethnic identity

Information collected using the CSS application has previously been provided.^17^ Briefly, at download and study enrollment, participants were asked to provide baseline demographic information, as well as details on suspected risk factors or relevant comorbidities (**Table 1**). They were asked with which race and/or ethnicity they self-identified based on standardized categories from the National Institutes of Health (NIH) in the U.S.^21^ and the Office for National Statistics in the U.K. (**Table S2**).^22^ In the U.S., Hispanic classification was defined as any race of Hispanic or Latino ancestry. Non-Hispanic categories were defined as each respective race not of Hispanic or Latino ancestry. Individuals who identified their race or ethnicity as “Other” were offered an opportunity to provide a free-text entry. Those who identified as “Mixed Race” or selected more than one race were categorized as “More than one race”. “Native Hawaiian and Pacific Islanders” were classified as Asian. Due to limited sample sizes, “American Indian or Alaskan Natives” were categorized as “Other”, and “Other” and “More than one race” were combined. In the U.K., individuals were asked whether they identified as “Chinese” or “Asian/British Asian”, which offered the following examples, but did not specifically ask about other racial identities from the Asian continent: “Indian, Pakistani, Bangladeshi, Other”. Responses were then aggregated in a manner consistent with prior analyses.^23^ We excluded individuals who selected “Prefer not to say” as their response or did not answer these questions.

**Table 2.** Vaccine hesitancy by race and ethnicity according to country of enrollment

|  | United States |  |  |  |  |
| --- | --- | --- | --- | --- | --- |
|  | White | Black | Hispanic | Asian | More than one/other |
| **Number hesitant or unsure / total** | 4715/64144 | 611/2179 | 505/3235 | 309/3089 | 166/1003 |
| **Age-adjusted OR (95% CI)<sup>1</sup>** | 1.0 (ref.) | 3.84 (3.51 to 4.21) | 1.69 (1.53 to 1.86) | 1.22 (1.03 to 1.38) | 2.14 (1.82 to 2.52) |
| **Multivariable-adjusted OR (95% CI)<sup>2</sup>** | 1.0 (ref.) | 3.68 (3.35 to 4.05) | 1.66 (1.50 to 1.84) | 1.33 (1.17 to 1.50) | 2.13 (1.80 to 2.52) |
| **Multivariable-adjusted OR (95% CI)<sup>3</sup>** | 1.0 (ref.) | 3.15 (2.86 to 3.47) | 1.42 (1.28 to 1.58) | 1.34 (1.18 to 1.52) | 2.02 (1.70 to 2.39) |

|  | United Kingdom |  |  |  |  |
| --- | --- | --- | --- | --- | --- |
|  | White | Black | South Asian | Middle East/East Asian | More than one/other |
| **Number hesitant or unsure / total** | 567734/1110544 | 1616/8787 | 1487/15199 | 771/6946 | 1270/13512 |
| **Age-adjusted OR (95% CI)<sup>1</sup>** | 1.0 (ref.) | 3.00 (2.86 to 3.16) | 1.59 (1.51 to 1.67) | 1.83 (1.70 to 1.97) | 1.43 (1.36 to 1.52) |
| **Multivariable-adjusted OR (95% CI)<sup>2</sup>** | 1.0 (ref.) | 2.96 (2.82 to 3.12) | 1.65 (1.56 to 1.73) | 1.84 (1.71 to 1.97) | 1.42 (1.34 to 1.50) |
| **Multivariable-adjusted OR (95% CI)<sup>3</sup>** | 1.0 (ref.) | 2.84 (2.69 to 2.99) | 1.66 (1.57 to 1.76) | 1.84 (1.7 to 1.98) | 1.48 (1.39 to 1.57) |

<sup>1</sup>Conditioned upon age and date of study entry
<sup>2</sup>Additional conditioning upon sex and adjustment for personal history of diabetes, heart disease, lung disease, kidney disease, current smoking status, body mass index, and prior reported history of COVID-19 infection
<sup>3</sup>Additional adjustment for frontline healthcare worker status, region, and education and income at the community level
Abbreviations: CI (confidence interval), OR (odds ratio)

### Community-level sociodemographic factors

Participants who elected to share information on their zip code (U.S./U.K.) or Lower Layer Super Output Area (LSOA, U.K.) of residence were assigned community-level socioeconomic measures. Socioeconomic measures were generated using established metrics derived from aggregated census data: proportion of individuals aged ≥25 years old with a Bachelor’s degree and median annual income (U.S.)^24^ and the education and income measures from the U.K.’s Indices of Multiple Deprivation (deciles, IMD),^25^ respectively. U.S. zip codes were further categorized into broader regions using U.S. Census Bureau criteria^26–28^ (American Northeast, Midwest, South, and West) and LSOAs were linked to one of four countries in the U.K. (England, Scotland, Northern Ireland, and Wales).^24^

### Ascertainment of other covariates and exposures

We collected information on age (years), sex at birth (male, female, or other), weight (kg) and height (meters) were used to calculate body mass index (BMI, <18.5, 18.5-24.9, 25-29.9, and ≥30 kg/m^2^), prior history of diabetes, heart disease, lung disease, kidney disease, or active malignancy (each yes/no), smoking history (current/prior vs. never), and frontline HCW status (yes/no). We longitudinally ascertained whether they had ever tested positive for COVID-19 (yes/no). To ascertain the validity of self-reported COVID Symptom Study information, we performed a validation study among a random subset of 235 users who reported a COVID-19 test from whom we requested photographs of their COVID-19 test reports. Study staff blinded to participant-provided information found excellent agreement between self-report and test reports with 88% sensitivity and 94% specificity.

### Statistical analysis

For missing data, imputation replaced no more than 5% of missing values for a given metadatum with numeric values replaced with the median and categorical variables imputed using the mode. Complete case analyses without imputation were performed to ensure robustness of findings and to ensure correction of missingness did not materially alter risk estimates (data not shown). LHN, ADJ, DAD, and ATC had access to raw data. LHN performed data analysis, and the corresponding author had full access to data and the final responsibility to submit for publication. Based on the size of the assembled study population and the empirically observed vaccine hesitancy of 6%, we had 80% power to detect a minimum OR of 1.12 and 1.07, respectively, in the U.S. and the U.K. for risk of (or protection from) reporting vaccine hesitancy among non-White compared to White participants.

To investigate determinants of COVID-19 vaccine hesitancy and uptake, we performed multivariable logistic regression to estimate odds ratios (OR) and their 95% confidence intervals (CIs) conditioned upon age, sex, and date of study entry adjusting for history of diabetes, heart disease, lung disease, kidney disease, cancer, current/prior smoking status, BMI, prior history of COVID-19 infection, occupation as frontline HCW, geographic region (U.S.)/country (U.K.), and sociodemographic factors based on community-level measures of educational and financial deprivation. We performed stratified analyses among frontline HCWs and the general community, and both lower and higher community-level educational attainment and financial deprivation, respectively. Formal tests for interaction were assessed using the Wald test in models with country-by-race/ethnicity interaction terms. Finally, we reported the prevalence of localized injection-site symptoms among vaccinated participants. Two-sided *p*-values were considered statistically significant. All statistical analyses were performed using R 4.0.3 (Vienna, Austria) and packages from the Bioconductor 3.12 release.

## Results

### Study population

From March 24, 2020 to February 16, 2021, we enrolled a total of 4,797,306 individuals (*n*=370,282 U.S. participants and *n*=4,427,024 U.K participants), of whom 1,894,940 individuals were active (logged at least one entry) as of Dec 24, 2020 (i.e., two weeks prior to the initial vaccine questionnaire). After excluding participants who did not provide their racial/ethnic information and restricting to those who responded to at least one vaccine questionnaires, a final analytic cohort of 1,341,682 individuals remained; **Fig S1**).

In the U.S., White participants tended to be older and reside in communities with higher income and educational attainment compared to Black or Hispanic individuals (**Table 1**). Black and Hispanic individuals more frequently reported being a frontline HCW and having previously been infected with SARS-CoV-2. Similar trends were observed in the U.K.

### Vaccine hesitancy among racial/ethnic minorities

Among 1,228,638 individuals who reported vaccine willingness, 91% of the 73,650 U.S. participants and 95% of the 1,154,988 U.K. participants were willing to accept a COVID-19 vaccine if offered (**Table S3**). In the U.S., those who were hesitant (unwilling/unsure) tended to be younger, female, less likely to have had heart disease or cancer, and more likely to live in communities with lower educational attainment and median incomes. Among frontline HCWs, 6% were unwilling to pursue vaccination and 11% were unsure, compared to 2% and 7% across the entire U.S. study population. Similar (younger) age distributions, burden of chronic disease, proportion of frontline HCWs, and rates of prior SARS-CoV-2 infection were observed among U.K. participants, though U.K. HCWs generally reported greater vaccine willingness.

**Table 3.** Vaccine uptake by race and ethnicity according to country of enrollment

**A. Vaccine uptake among all participants**
|  | United States |  |  |  |  |
| --- | --- | --- | --- | --- | --- |
|  | White | Black | Hispanic | Asian | More than one/other |
| **Number receiving a vaccine / total** | 15341/64144 | 362/2179 | 519/3235 | 716/3089 | 202/1003 |
| **Age-adjusted OR (95% CI)<sup>1</sup>** | 1.0 (ref.) | 0.76 (0.68 to 0.84) | 1.00 (0.92 to 1.10) | 1.10 (1.02 to 1.19) | 0.97 (0.84 to 1.11) |
| **Multivariable-adjusted OR (95% CI)<sup>2</sup>** | 1.0 (ref.) | 0.76 (0.68 to 0.85) | 1.01 (0.93 to 1.11) | 1.07 (0.99 to 1.15) | 0.96 (0.83 to 1.10) |
| **Multivariable-adjusted OR (95% CI)<sup>3</sup>** | 1.0 (ref.) | 0.71 (0.64 to 0.79) | 0.93 (0.84 to 1.02) | 1.00 (0.93 to 1.09) | 0.94 (0.81 to 1.08) |

|  | United Kingdom |  |  |  |  |
| --- | --- | --- | --- | --- | --- |
|  | White | Black | South Asian | Middle East/East Asian | More than one/other |
| **Number receiving a vaccine / total** | 171453/1110544 | 1022/8787 | 2339/15199 | 922/6946 | 1506/13512 |
| **Age-adjusted OR (95% CI)<sup>1</sup>** | 1.0 (ref.) | 1.13 (1.06 to 1.20) | 1.34 (1.29 to 1.40) | 1.10 (1.03 to 1.18) | 1.05 (1.00 to 1.11) |
| **Multivariable-adjusted OR (95% CI)<sup>2</sup>** | 1.0 (ref.) | 1.12 (1.06 to 1.19) | 1.31 (1.25 to 1.36) | 1.09 (1.02 to 1.16) | 1.04 (0.98 to 1.09) |
| **Multivariable-adjusted OR (95% CI)<sup>3</sup>** | 1.0 (ref.) | 0.98 (0.92 to 1.04) | 1.18 (1.13 to 1.23) | 1.01 (0.94 to 1.08) | 0.99 (0.93 to 1.04) |

<sup>1</sup>Conditioned upon age and date of study entry
<sup>2</sup>Additional conditioning upon sex and adjustment for personal history of diabetes, heart disease, lung disease, kidney disease, current smoking status, body mass index, and prior reported history of COVID-19 infection
<sup>3</sup>Additional adjustment for frontline healthcare worker status, region, and education and income at the community level
Abbreviations: CI (confidence interval), OR (odds ratio). Data shown through February 1, 2021.

B. Vaccine uptake among vaccine willing
|  | United States |  |  |  |  |
| --- | --- | --- | --- | --- | --- |
|  | White | Black | Hispanic | Asian | More than one/other |
| **Number receiving a vaccine / total** | 15062/59429 | 328/1568 | 499/2730 | 681/2780 | 192/837 |
| **Age-adjusted OR (95% CI)<sup>1</sup>** | 1.0 (ref.) | 0.88 (0.78 to 0.98) | 1.04 (0.95 to 1.14) | 1.11 (1.03 to 1.20) | 1.03 (0.89 to 1.19) |
| **Multivariable-adjusted OR (95% CI)<sup>2</sup>** | 1.0 (ref.) | 0.88 (0.78 to 0.98) | 1.05 (0.96 to 1.15) | 1.08 (1.00 to 1.17) | 1.02 (0.88 to 1.17) |
| **Multivariable-adjusted OR (95% CI)<sup>3</sup>** | 1.0 (ref.) | 0.82 (0.73 to 0.92) | 0.95 (0.86 to 1.04) | 1.01 (0.93 to 1.09) | 1.00 (0.86 to 1.16) |

|  | United Kingdom |  |  |  |  |
| --- | --- | --- | --- | --- | --- |
|  | White | Black | South Asian | Middle East/East Asian | More than one/other |
| **Number receiving a vaccine / total** | 168369/1053810 | 951/7171 | 2255/13712 | 884/6175 | 1469/12242 |
| **Age-adjusted OR (95% CI)<sup>1</sup>** | 1.0 (ref.) | 1.22 (1.14 to 1.30) | 1.37 (1.31 to 1.42) | 1.13 (1.06 to 1.21) | 1.09 (1.03 to 1.15) |
| **Multivariable-adjusted OR (95% CI)<sup>2</sup>** | 1.0 (ref.) | 1.22 (1.14 to 1.30) | 1.33 (1.28 to 1.39) | 1.12 (1.05 to 1.20) | 1.07 (1.01 to 1.13) |
| **Multivariable-adjusted OR (95% CI)<sup>3</sup>** | 1.0 (ref.) | 1.07 (1.00 to 1.14) | 1.21 (1.16 to 1.26) | 1.04 (0.97 to 1.11) | 1.02 (0.97 to 1.08) |

<sup>1</sup>Conditioned upon age and date of study entry
<sup>2</sup>Additional conditioning upon sex and adjustment for personal history of diabetes, heart disease, lung disease, kidney disease, current smoking status, body mass index, and prior reported history of COVID-19 infection
<sup>3</sup>Additional adjustment for frontline healthcare worker status, region, and education and income at the community level
Abbreviations: CI (confidence interval), OR (odds ratio). Data shown through February 1, 2021.

In the U.S. and U.K., racial/ethnic minorities were more likely to report being unsure or unwilling to undergo vaccination. In the U.S., compared to White, the age-adjusted ORs for vaccine hesitancy were 3.84 (95% CI: 3.51 to 4.21) for Black, 1.69 (95% CI: 1.53 to 1.86) for Hispanic, and 1.22 (95% CI: 1.03 to 1.38) for Asian individuals, and 2.14 (95% CI: 1.82 to 2.52) for those who reported other or more than one race (**Table 2**). Additional adjustment for relevant covariates did not materially alter these risk estimates. Similar degrees of hesitancy were observed among racial and ethnic minorities in the U.K., which was most striking among Black and Hispanic individuals (**Table 2**).

In the U.S., we observed regional differences in willingness to be vaccinated with greater hesitancy in the South (**Table S4**). In the U.K., compared to participants in England, the age-adjusted ORs for vaccine hesitancy were 1.38 (1.25 to 1.51) for participants from Northern Ireland and 1.10 (1.06 to 1.15) for Wales. These were not substantially altered after additional adjustment.

**Table 4.** Vaccine uptake by race and ethnicity according to country of enrollment by participant subgroup

**A. Vaccine uptake among frontline healthcare workers and the general community**
|  | United States |  |  |  |  |
| --- | --- | --- | --- | --- | --- |
|  | White | Black | Hispanic | Asian | More than one/other |
| **<u>Frontline healthcare workers</u>** |  |  |  |  |  |
| **Number receiving a vaccine / total** | 827/1906 | 28/112 | 41/136 | 41/89 | 18/44 |
| **Age-adjusted OR (95% CI)<sup>1</sup>** | 1.0 (ref.) | 0.59 (0.38 to 0.92) | 0.85 (0.57 to 1.27) | 1.00 (0.64 to 1.55) | 1.01 (0.54 to 1.90) |
| **Multivariable-adjusted OR (95% CI)<sup>2</sup>** | 1.0 (ref.) | 0.63 (0.39 to 1.03) | 0.81 (0.52 to 1.25) | 1.11 (0.68 to 1.80) | 1.02 (0.50 to 2.06) |
| **Multivariable-adjusted OR (95% CI)<sup>3</sup>** | 1.0 (ref.) | 0.67 (0.40 to 1.11) | 0.84 (0.53 to 1.33) | 1.01 (0.62 to 1.66) | 1.04 (0.51 to 2.10) |
| **<u>General community</u>** |  |  |  |  |  |
| **Number receiving a vaccine / total** | 14514/62238 | 334/2067 | 478/3099 | 675/3000 | 184/959 |
| **Age-adjusted OR (95% CI)<sup>1</sup>** | 1.0 (ref.) | 0.75 (0.68 to 0.84) | 1.01 (0.92 to 1.11) | 1.10 (1.01 to 1.18) | 0.95 (0.82 to 1.10) |
| **Multivariable-adjusted OR (95% CI)<sup>2</sup>** | 1.0 (ref.) | 0.76 (0.68 to 0.85) | 1.02 (0.93 to 1.12) | 1.07 (0.98 to 1.15) | 0.94 (0.81 to 1.09) |
| **Multivariable-adjusted OR (95% CI)<sup>3</sup>** | 1.0 (ref.) | 0.73 (0.65 to 0.82) | 0.93 (0.85 to 1.03) | 0.99 (0.91 to 1.08) | 0.91 (0.79 to 1.06) |

<sup>1</sup>Conditioned upon age and date of study entry
<sup>2</sup>Additional conditioning upon sex and adjustment for personal history of diabetes, heart disease, lung disease, kidney disease, current smoking status, body mass index, and prior reported history of COVID-19 infection
<sup>3</sup>Additional adjustment for frontline healthcare worker status, region, and education and income at the community level (except in models for a given strata)
Abbreviations: CI (confidence interval), OR (odds ratio). Data shown through February 1, 2021.

|  | United Kingdom |  |  |  |  |
| --- | --- | --- | --- | --- | --- |
|  | White | Black | South Asian | Middle East/East Asian | More than one/other |
| **Frontline healthcare workers** |  |  |  |  |  |
| **Number receiving a vaccine / total** | 14955/25449 | 158/373 | 313/579 | 106/204 | 152/320 |
| **Age-adjusted OR (95% CI)<sup>1</sup>** | 1.0 (ref.) | 0.70 (0.59 to 0.82) | 0.94 (0.84 to 1.06) | 0.87 (0.72 to 1.06) | 0.79 (0.67 to 0.93) |
| **Multivariable-adjusted OR (95% CI)<sup>2</sup>** | 1.0 (ref.) | 0.71 (0.6 to 0.84) | 0.97 (0.86 to 1.10) | 0.85 (0.70 to 1.05) | 0.81 (0.68 to 0.96) |
| **Multivariable-adjusted OR (95% CI)<sup>3</sup>** | 1.0 (ref.) | 0.71 (0.6 to 0.85) | 0.95 (0.84 to 1.09) | 0.85 (0.68 to 1.05) | 0.78 (0.65 to 0.94) |
| **General community** |  |  |  |  |  |
| **Number receiving a vaccine / total** | 156498/1085095 | 864/8414 | 2026/14620 | 816/6742 | 1354/13192 |
| **Age-adjusted OR (95% CI)<sup>1</sup>** | 1.0 (ref.) | 1.13 (1.06 to 1.21) | 1.34 (1.28 to 1.4) | 1.11 (1.04 to 1.19) | 1.10 (1.04 to 1.16) |
| **Multivariable-adjusted OR (95% CI)<sup>2</sup>** | 1.0 (ref.) | 1.13 (1.05 to 1.21) | 1.30 (1.24 to 1.36) | 1.10 (1.02 to 1.18) | 1.08 (1.02 to 1.14) |
| **Multivariable-adjusted OR (95% CI)<sup>3</sup>** | 1.0 (ref.) | 1.05 (0.98 to 1.13) | 1.22 (1.17 to 1.28) | 1.05 (0.98 to 1.13) | 1.02 (0.97 to 1.08) |

<sup>1</sup>Conditioned upon age and date of study entry
<sup>2</sup>Additional conditioning upon sex and adjustment for personal history of diabetes, heart disease, lung disease, kidney disease, current smoking status, body mass index, and prior reported history of COVID-19 infection
<sup>3</sup>Additional adjustment for frontline healthcare worker status, region, and education and income at the community level (except in models for a given strata)
Abbreviations: CI (confidence interval), OR (odds ratio). Data shown through February 1, 2021.

B. Vaccine uptake by economic deprivation
|  | United States |  |  |  |  |
| --- | --- | --- | --- | --- | --- |
|  | White | Black | Hispanic | Asian | More than one/other |
| **<u>Lower income (Quartile 1)</u>** |  |  |  |  |  |
| **Number receiving a vaccine / total** | 3660/15207 | 156/937 | 152/1033 | 71/421 | 55/267 |
| **Age-adjusted OR (95% CI)<sup>1</sup>** | 1.0 (ref.) | 0.73 (0.62 to 0.86) | 0.99 (0.84 to 1.18) | 0.90 (0.71 to 1.15) | 1.13 (0.85 to 1.49) |
| **Multivariable-adjusted OR (95% CI)<sup>2</sup>** | 1.0 (ref.) | 0.75 (0.63 to 0.89) | 0.99 (0.83 to 1.19) | 0.89 (0.69 to 1.14) | 1.07 (0.80 to 1.43) |
| **Multivariable-adjusted OR (95% CI)<sup>3</sup>** | 1.0 (ref.) | 0.73 (0.61 to 0.86) | 0.94 (0.79 to 1.14) | 0.90 (0.70 to 1.16) | 1.08 (0.81 to 1.44) |
| **<u>Higher income (Quartile 4)</u>** |  |  |  |  |  |
| **Number receiving a vaccine / total** | 3645/15599 | 54/302 | 115/630 | 244/1089 | 53/225 |
| **Age-adjusted OR (95% CI)<sup>1</sup>** | 1.0 (ref.) | 0.86 (0.65 to 1.13) | 1.13 (0.93 to 1.36) | 1.09 (0.95 to 1.24) | 1.13 (0.85 to 1.50) |
| **Multivariable-adjusted OR (95% CI)<sup>2</sup>** | 1.0 (ref.) | 0.85 (0.64 to 1.13) | 1.15 (0.94 to 1.39) | 1.08 (0.94 to 1.24) | 1.12 (0.84 to 1.49) |
| **Multivariable-adjusted OR (95% CI)<sup>3</sup>** | 1.0 (ref.) | 0.83 (0.62 to 1.11) | 1.05 (0.86 to 1.28) | 0.97 (0.84 to 1.12) | 1.09 (0.81 to 1.45) |

<sup>1</sup>Conditioned upon age and date of study entry
<sup>2</sup>Additional conditioning upon sex and adjustment for personal history of diabetes, heart disease, lung disease, kidney disease, current smoking status, body mass index, and prior reported history of COVID-19 infection
<sup>3</sup>Additional adjustment for frontline healthcare worker status, region, and education and income at the community level (except in models for a given strata)
Abbreviations: CI (confidence interval), OR (odds ratio). Data shown through February 1, 2021.

|  | United Kingdom |  |  |  |  |
| --- | --- | --- | --- | --- | --- |
|  | White | Black | South Asian | Middle East/East Asian | More than one/other |
| **Lower income (Quartile 1)** |  |  |  |  |  |
| **Number receiving a vaccine / total** | 41761/283212 | 455/3904 | 745/5377 | 289/2260 | 459/4524 |
| **Age-adjusted OR (95% CI)<sup>1</sup>** | 1.0 (ref.) | 1.12 (1.02 to 1.23) | 1.24 (1.15 to 1.33) | 1.15 (1.02 to 1.29) | 0.97 (0.88 to 1.06) |
| **Multivariable-adjusted OR (95% CI)<sup>2</sup>** | 1.0 (ref.) | 1.11 (1.01 to 1.22) | 1.21 (1.12 to 1.30) | 1.15 (1.02 to 1.29) | 0.95 (0.87 to 1.04) |
| **Multivariable-adjusted OR (95% CI)<sup>3</sup>** | 1.0 (ref.) | 0.97 (0.89 to 1.07) | 1.12 (1.04 to 1.21) | 1.05 (0.94 to 1.18) | 0.90 (0.82 to 0.99) |
| **Higher income (Quartile 4)** |  |  |  |  |  |
| **Number receiving a vaccine / total** | 29993/192710 | 111/863 | 345/2250 | 136/1063 | 224/2018 |
| **Age-adjusted OR (95% CI)<sup>1</sup>** | 1.0 (ref.) | 1.18 (0.98 to 1.42) | 1.32 (1.19 to 1.47) | 0.99 (0.83 to 1.17) | 1.04 (0.91 to 1.19) |
| **Multivariable-adjusted OR (95% CI)<sup>2</sup>** | 1.0 (ref.) | 1.20 (1.00 to 1.45) | 1.29 (1.16 to 1.43) | 0.97 (0.81 to 1.14) | 1.03 (0.90 to 1.18) |
| **Multivariable-adjusted OR (95% CI)<sup>3</sup>** | 1.0 (ref.) | 1.08 (0.90 to 1.31) | 1.19 (1.07 to 1.33) | 0.91 (0.77 to 1.08) | 1.01 (0.88 to 1.15) |

<sup>1</sup>Conditioned upon age and date of study entry
<sup>2</sup>Additional conditioning upon sex and adjustment for personal history of diabetes, heart disease, lung disease, kidney disease, current smoking status, body mass index, and prior reported history of COVID-19 infection
<sup>3</sup>Additional adjustment for frontline healthcare worker status, region, and education and income at the community level (except in models for a given strata)
Abbreviations: CI (confidence interval), OR (odds ratio). Data shown through February 1, 2021.

C. Vaccine uptake by educational attainment
|  | United States |  |  |  |  |
| --- | --- | --- | --- | --- | --- |
|  | White | Black | Hispanic | Asian | More than one/other |
| **Lower educational attainment (Quartile 1)** |  |  |  |  |  |
| **Number receiving a vaccine / total** | 3441/14875 | 141/926 | 189/1221 | 165/610 | 55/304 |
| **Age-adjusted OR (95% CI)<sup>1</sup>** | 1.0 (ref.) | 0.69 (0.58 to 0.82) | 1.11 (0.95 to 1.30) | 1.18 (1.01 to 1.39) | 0.91 (0.69 to 1.21) |
| **Multivariable-adjusted OR (95% CI)<sup>2</sup>** | 1.0 (ref.) | 0.70 (0.58 to 0.83) | 1.15 (0.98 to 1.36) | 1.14 (0.96 to 1.35) | 0.87 (0.66 to 1.16) |
| **Multivariable-adjusted OR (95% CI)<sup>3</sup>** | 1.0 (ref.) | 0.65 (0.54 to 0.78) | 1.06 (0.90 to 1.26) | 1.10 (0.92 to 1.31) | 0.86 (0.64 to 1.14) |
| **Higher educational attainment (Quartile 4)** |  |  |  |  |  |
| **Number receiving a vaccine / total** | 3808/16031 | 56/278 | 88/528 | 155/800 | 46/214 |
| **Age-adjusted OR (95% CI)<sup>1</sup>** | 1.0 (ref.) | 0.96 (0.73 to 1.26) | 1.03 (0.83 to 1.28) | 1.27 (1.07 to 1.50) | 1.06 (0.78 to 1.44) |
| **Multivariable-adjusted OR (95% CI)<sup>2</sup>** | 1.0 (ref.) | 0.97 (0.73 to 1.28) | 1.05 (0.84 to 1.32) | 1.28 (1.08 to 1.52) | 1.06 (0.78 to 1.45) |
| **Multivariable-adjusted OR (95% CI)<sup>3</sup>** | 1.0 (ref.) | 0.93 (0.70 to 1.24) | 0.95 (0.75 to 1.19) | 1.19 (1.00 to 1.42) | 1.09 (0.80 to 1.50) |

<sup>1</sup>Conditioned upon age and date of study entry
<sup>2</sup>Additional conditioning upon sex and adjustment for personal history of diabetes, heart disease, lung disease, kidney disease, current smoking status, body mass index, and prior reported history of COVID-19 infection
<sup>3</sup>Additional adjustment for frontline healthcare worker status, region, and education and income at the community level (except in models for a given strata)
Abbreviations: CI (confidence interval), OR (odds ratio). Data shown through February 1, 2021.

|  | United Kingdom |  |  |  |  |
| --- | --- | --- | --- | --- | --- |
|  | White | Black | South Asian | Middle East/East Asian | More than one/other |
| **<u>Lower educational attainment (Quartile 1)</u>** |  |  |  |  |  |
| **Number receiving a vaccine / total** | 39534/273555 | 354/2822 | 439/3422 | 178/1324 | 289/2881 |
| **Age-adjusted OR (95% CI)<sup>1</sup>** | 1.0 (ref.) | 1.22 (1.10 to 1.36) | 1.14 (1.03 to 1.25) | 1.16 (1.00 to 1.35) | 0.94 (0.84 to 1.05) |
| **Multivariable-adjusted OR (95% CI)<sup>2</sup>** | 1.0 (ref.) | 1.21 (1.09 to 1.34) | 1.13 (1.03 to 1.24) | 1.18 (1.02 to 1.37) | 0.93 (0.83 to 1.04) |
| **Multivariable-adjusted OR (95% CI)<sup>3</sup>** | 1.0 (ref.) | 1.08 (0.97 to 1.20) | 1.06 (0.97 to 1.17) | 1.09 (0.94 to 1.27) | 0.94 (0.83 to 1.05) |
| **<u>Higher educational attainment (Quartile 4)</u>** |  |  |  |  |  |
| **Number receiving a vaccine / total** | 35969/217167 | 153/1259 | 503/3215 | 220/1632 | 346/3110 |
| **Age-adjusted OR (95% CI)<sup>1</sup>** | 1.0 (ref.) | 1.07 (0.91 to 1.25) | 1.30 (1.19 to 1.43) | 1.05 (0.92 to 1.20) | 1.03 (0.93 to 1.14) |
| **Multivariable-adjusted OR (95% CI)<sup>2</sup>** | 1.0 (ref.) | 1.06 (0.90 to 1.24) | 1.27 (1.16 to 1.39) | 1.02 (0.89 to 1.17) | 1.01 (0.91 to 1.13) |
| **Multivariable-adjusted OR (95% CI)<sup>3</sup>** | 1.0 (ref.) | 0.96 (0.81 to 1.12) | 1.13 (1.03 to 1.23) | 0.93 (0.81 to 1.06) | 0.96 (0.86 to 1.07) |

<sup>1</sup>Conditioned upon age and date of study entry
<sup>2</sup>Additional conditioning upon sex and adjustment for personal history of diabetes, heart disease, lung disease, kidney disease, current smoking status, body mass index, and prior reported history of COVID-19 infection
<sup>3</sup>Additional adjustment for frontline healthcare worker status, region, and education and income at the community level (except in models for a given strata)
Abbreviations: CI (confidence interval), OR (odds ratio). Data shown through February 1, 2021.

When exploring the specific reasons for reluctance, the most frequently indicated concerns among all race and ethnicities related to long-term side effects (50-57%) and adverse reactions (45-54%). Additionally, Black and Hispanic individuals cited a lack of knowledge about the vaccine (45-51%) at a higher rate than White individuals (37-42%; **Table S5**).

### Racial/ethnic disparities in COVID-19 vaccine uptake

Based on eligibility in the initial phase of mass vaccinations, as expected, vaccinated participants through February 1, 2021 tended to be older, had greater comorbidities, and were considerably more likely to be frontline HCWs (**Table S6**). In the U.S., Black individuals were less likely to be vaccinated than White participants (OR 0.71, 95% CI: 0.64 to 0.79), even after adjusting for age, region, comorbidities, and occupation as a HCW (**Table 3A**). In a subgroup analysis, these associations persisted even when we limited analysis to individuals who reported vaccine-willingness (**Table 3B**). In contrast, in the U.K, non-White participants reported higher vaccination rates than White individuals, including Black, South Asian, and Middle East/East Asian persons, though adjustment for personal and community risk factors attenuated these results.

The disparity in vaccine uptake among Black individuals compared with White individuals differed significantly by country (P_heterogeneity_<0.001). When compared to White individuals within their respective countries, Black individuals were considerably less likely to be vaccinated in the U.S. (**Fig. 1**). Compared to the Northeast, vaccine uptake was comparatively greater in other parts of the U.S. In the U.K., England appeared to have greater vaccine uptake compared to other countries where increased vaccine hesitancy has been documented (**Table S7**).^30, 31^

**Figure 1.**
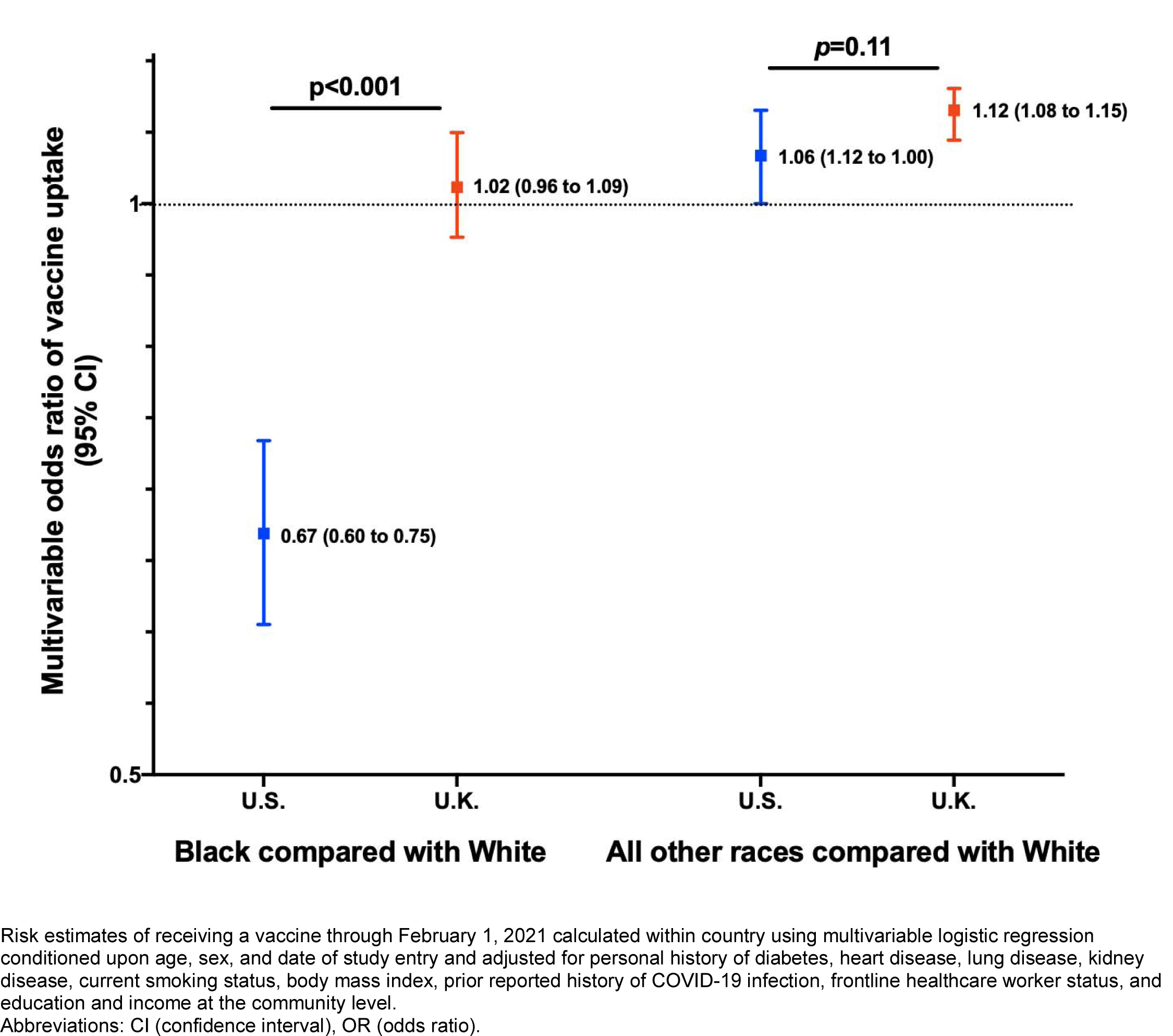
Disparity in vaccine uptake by race and ethnicity according to country of enrollment Risk estimates of receiving a vaccine through February 1, 2021 calculated within country using multivariable logistic regression conditioned upon age, sex, and date of study entry and adjusted for personal history of diabetes, heart disease, lung disease, kidney disease, current smoking status, body mass index, prior reported history of COVID-19 infection, frontline healthcare worker status, and education and income at the community level. Abbreviations: CI (confidence interval), OR (odds ratio).

Lower vaccine uptake among Black persons in the U.S. were comparable among specific sociodemographic groups, including frontline HCWs (**Table 4A**) and individuals living in communities with lower educational attainment (**Table 4C**). Notably, in the U.K., Black frontline HCWs had lower vaccine uptake than their White counterparts (**Table 4B**). Finally, no consistent differences were observed in local vaccine reactions according to race and ethnicity (**Table S8**).

## Discussion

Among 1,341,682 participants in the U.S. and U.K., we observed increased COVID-19 vaccine hesitancy among racial/ethnic minority participants largely driven by concerns about long-term safety and fear of adverse reactions. Through the early phase of the vaccination campaign (February 1, 2021), we revealed significant racial and ethnic disparities in uptake in the U.S., but not the U.K., even among those willing to receive a vaccine, suggesting issues related to access in the U.S. Interestingly, we observed a higher than anticipated rate of vaccine hesitancy among frontline HCWs, perhaps due to their substantially higher rate of prior COVID-19 infection and heightened concern about the relative lack of initial safety data.

Our findings of greater vaccine hesitancy among minority populations has been previously shown in smaller, more limited investigations.^32^ Deep-rooted and ongoing mistrust of the medical system among people of color^33^ and a lack of diverse representation in clinical trials^34^ likely play a substantial role in explaining this hesitancy. The speed with which vaccines were approved has raised suspicions over whether regulatory standards meant to protect vulnerable populations were relaxed for expediency.^35^ Moreover, racial and ethnic minorities who have already borne the disproportionate brunt of the pandemic^36^ may be taking a more cautious approach to new vaccines, particularly with our current incomplete understanding of post-infectious immunity and potential adverse effects. Reassuringly, our data did not reveal significant differences in localized injection-site reactions by race/ethnicity. Prior work specifically examining attitudes toward COVID-19 vaccines further support our findings. A recent randomized controlled trial demonstrated that COVID-19 vaccine misinformation significantly reduced vaccination intent in the U.K. and U.S.^33^ Notably, differences in susceptibility and receptiveness were observed across sociodemographic groups.

Our findings demonstrating lower vaccine uptake among communities of color have been shown in other studies. A recent study of U.K. HCWs showed substantially lower vaccine uptake among racial and ethnic minorities.^34^ Our results significantly extend these data by concurrently examining vaccine hesitancy and receipt within the same participants from sizable community-based samples in two countries. Importantly, we found that even among the U.S. vaccine-willing population, Black individuals were less likely to receive a vaccine, whereas in the U.K., no consistent disparities in vaccine uptake were observed.

The strengths of our study include the prospective population-scale enrollment of a diverse group of participants from two comparably afflicted nations using a common data collection instrument. With disparate approaches to COVID-19 vaccination campaigns and healthcare delivery in the U.S. and U.K., our multinational study design provided a unique opportunity to consider the degree to which structural inequities, public mistrust, and unequal care access could result in differences in vaccine willingness and uptake. Our use of a digital platform to rapidly collect this information on vaccine skepticism and usage provides real-time actionable insights to inform the public health response to an ongoing pandemic. Finally, extensive demographic information is generally not available in registry-level data or large-scale surveillance efforts, and we had an opportunity of uncommon breadth and depth to evaluate whether these established risk factors could influence vaccine attitudes and uptake.

We acknowledge several limitations. We relied primarily on volunteered information which may be subject to measurement and reporting bias. However, our validation study (**Methods**) demonstrates that self-reported information from the general population was accurately and faithfully reported. While our study had comparatively lower proportions of minorities, we enrolled relatively high absolute numbers for most groups. To maximize participation, greater detail on racial/ethnic self-identity could not be obtained, and our current categorizations may oversimplify or incompletely characterize the different lived experiences of minority participants navigating the healthcare system. Despite more than 80% of U.S. adults adopting smartphones,^35^ we acknowledge that our data collection may have comparatively lower penetrance among certain socioeconomic/age groups, though under-recruitment of more deprived or less technologically literate participants would likely have attenuated observed differences. In fact, the use of a mobile app for data collection allowed us to highlight racial/ethnic disparities in uptake that persisted despite access to technology. Finally, our cohort of study volunteers willing to share information about COVID-19 does not represent a random sampling of the U.S. and U.K. population and are likely enriched for individuals that are generally more accepting of vaccinations. Nonetheless, the significant differences we observed in vaccine hesitancy and uptake between racial groups remain internally valid and likely underestimate broader disparities within population samples that do not use a common data collection instrument.

In conclusion, we found significantly higher likelihood of COVID-19 vaccine hesitancy among racial and ethnic minority groups in the U.S. and the U.K., particularly among Black and Hispanic individuals, driven by concerns about adverse effects, supporting the need for targeted vaccine education from trusted messengers. Furthermore, in the U.S., a striking disparity in vaccination uptake among Black Americans was observed even among vaccine-willing individuals with similar access to mobile technology to White Americans. This difference, not found in the U.K., suggests that early disparities in U.S. vaccine uptake have been exacerbated by inequities in the fairness of prioritization and distribution of vaccines to minority communities. Given the relative lack of a national public health infrastructure in the U.S., our data highlight the potential value of a more centralized system of vaccine delivery to ensure equitable vaccine uptake. Taken together, these findings support the need to address long-standing systemic disparities to achieve the health equity required for population-scale immunity.

### Data Sharing

Data collected using the COVID Symptom Study smartphone application are being shared with other health researchers through the U.K. National Health Service-funded Health Data Research UK (HDRUK) and Secure Anonymised Information Linkage consortium, housed in the U.K. Secure Research Platform (Swansea, UK). Anonymized data are available to be shared with researchers according to their protocols in the public interest (https://web.www.healthdatagateway.org/dataset/fddcb382-3051-4394-8436-b92295f14259). U.S. investigators are encouraged to coordinate data requests through the Coronavirus Pandemic Epidemiology (COPE) Consortium (https://www.monganinstitute.org/cope-consortium).

### Declaration of Interests

LP, CH, SS, RD, and JW are employees of Zoe Global Ltd. TDS is a consultant to Zoe Global Ltd. DAD and ATC previously served as investigators on a clinical trial of diet and lifestyle using a separate smartphone application that was supported by Zoe Global Ltd.

## Funding

Zoe provided in kind support for all aspects of building, running and supporting the app and service to users worldwide. LHN is supported by the NIH/NIDDK NIH K23DK125838, American Gastroenterological Association Research Scholars Award, and Crohn’s and Colitis Foundation Career Development Award and Research Fellowship Award. DAD is supported by the NIH/NIDDK K01DK120742. LHN and DAD are supported by the American Gastroenterological Association-Takeda COVID-19 Rapid Response Research Award (AGA2021-5102). The Massachusetts Consortium on Pathogen Readiness (MassCPR) and Mark and Lisa Schwartz supported MGH investigators. CM is supported by the Chronic Disease Research Foundation. CM and TDS are supported by the Medical Research Council AimHy Project Grant (MR/M016560/1). ATC is the Stuart and Suzanne Steele MGH Research Scholar and Stand Up to Cancer scientist. Sponsors had no role in study design, analysis, and interpretation of data, report writing, and the decision to submit for publication.

## Data Availability

Data collected using the COVID Symptom Study smartphone application are being shared with other health researchers through the U.K. National Health Service-funded Health Data Research UK (HDRUK) and Secure Anonymised Information Linkage consortium, housed in the U.K. Secure Research Platform (Swansea, UK). Anonymized data are available to be shared with researchers according to their protocols in the public interest. U.S. investigators are encouraged to coordinate data requests through the Coronavirus Pandemic Epidemiology (COPE) Consortium.

---

**Figure S1.**
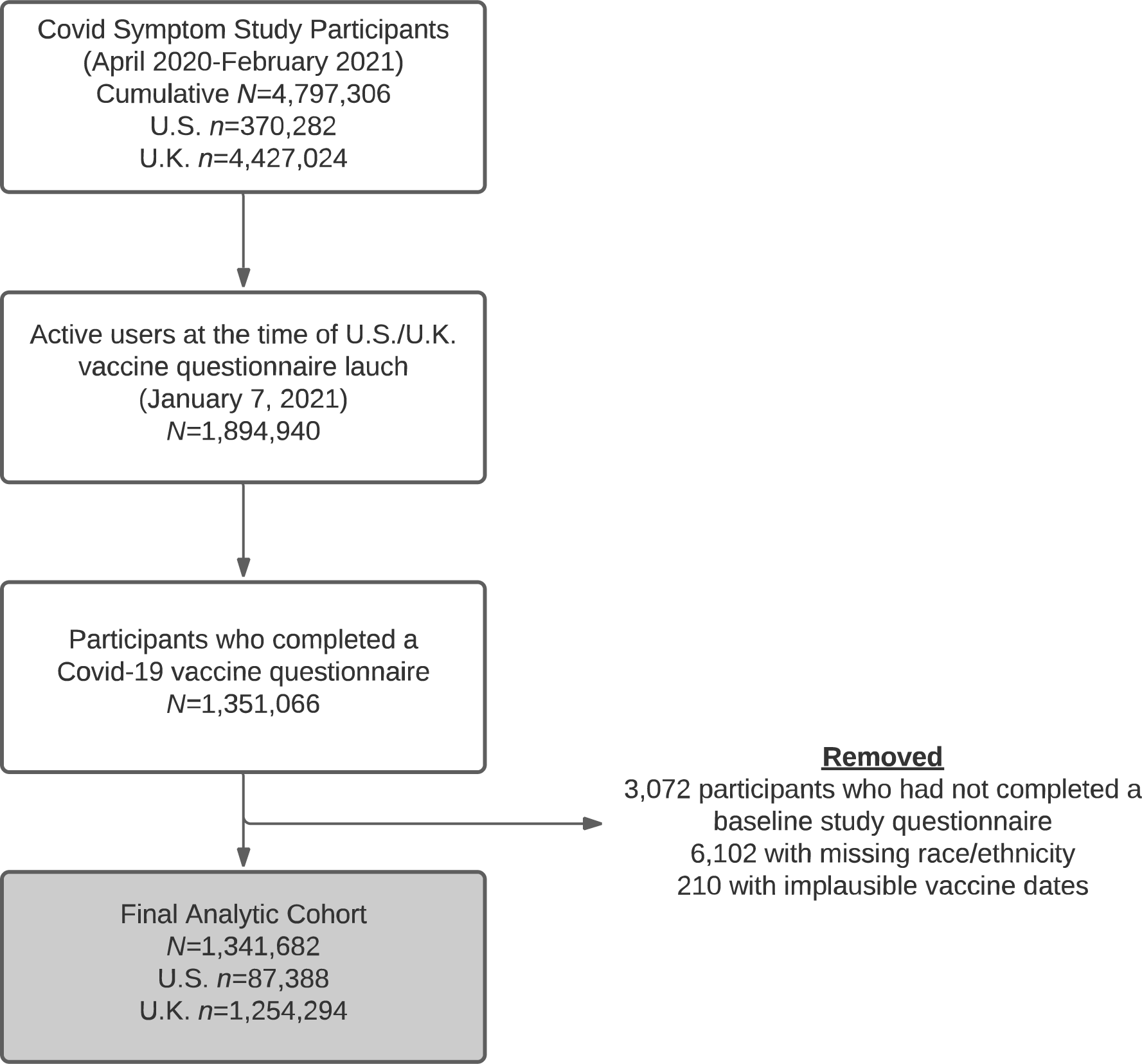
Study diagram

**Table S1.**
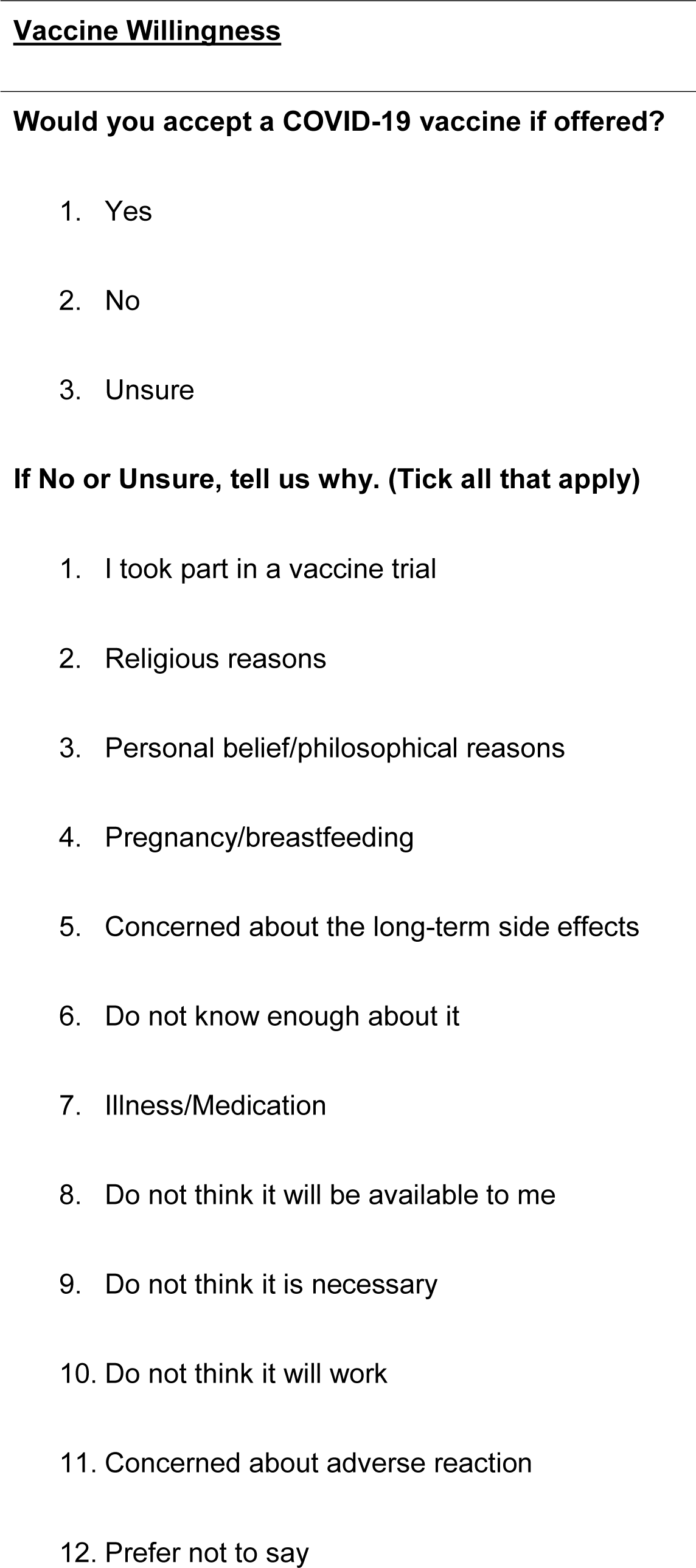
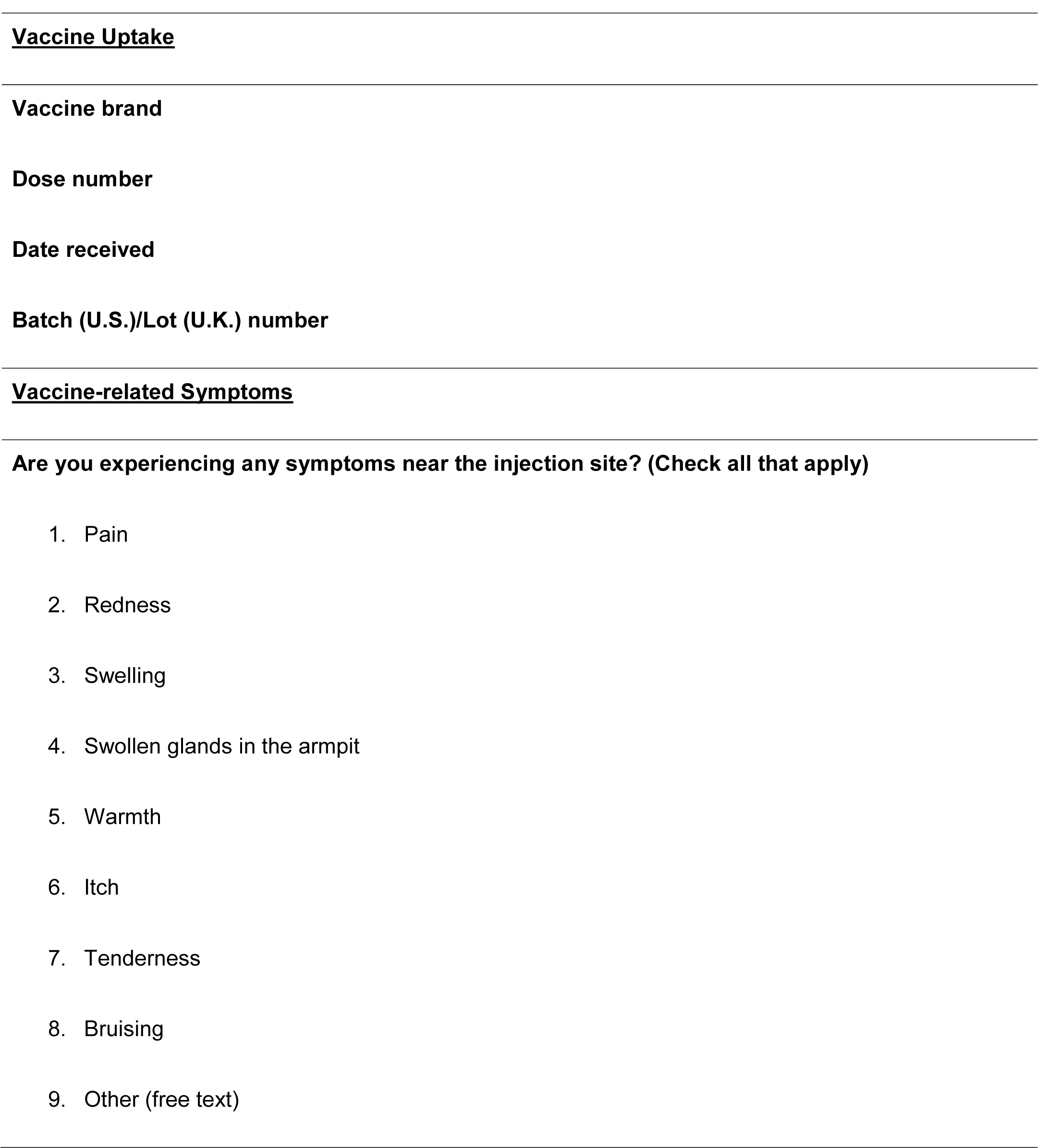
Vaccine-related questions through February 1, 2021.

**Table S2.**
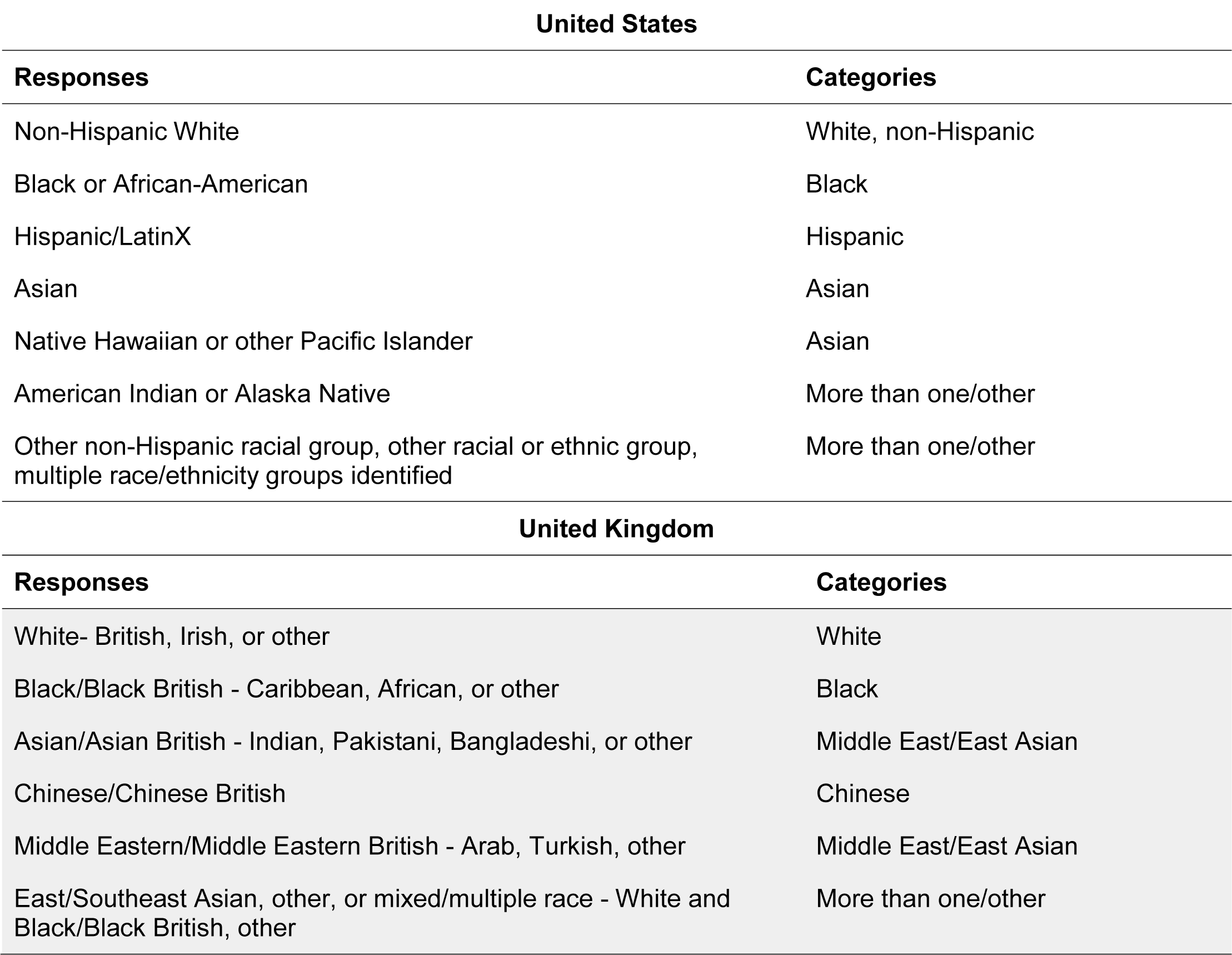
Race and ethnicity categories by country of enrollment.

**Table S3.** Baseline characteristics of study participants by vaccine willingness and country of enrollment.

|  | United States |  |  |
| --- | --- | --- | --- |
|  | Willing<br>(n=67344) | Unsure<br>(n=4943) | Unwilling<br>(n=1363) |
| **Age (years)** | 60.8 (15.5) | 54.7 (17.2) | 54.1 (16.8) |
| <25 | 2090 (3.1) | 317 (6.4) | 82 (6.0) |
| 25-34 | 3681 (5.5) | 486 (9.8) | 145 (10.6) |
| 35-44 | 5615 (8.3) | 582 (11.8) | 165 (12.1) |
| 45-54 | 7548 (11.2) | 783 (15.8) | 217 (15.9) |
| 55-64 | 13584 (20.2) | 1063 (21.5) | 320 (23.5) |
| ≥65 | 34826 (51.7) | 1712 (34.6) | 434 (31.8) |
| **Female sex** | 43181 (64.1) | 3597 (72.8) | 971 (71.2) |
| **Race/ethnicity** |  |  |  |
| White | 59429 (88.2) | 3708 (75.0) | 1007 (73.9) |
| Black | 1568 (2.3) | 468 (9.5) | 143 (10.5) |
| Hispanic | 2730 (4.1) | 385 (7.8) | 120 (8.8) |
| Asian | 2780 (4.1) | 262 (5.3) | 47 (3.4) |
| More than one / other | 837 (1.2) | 120 (2.4) | 46 (3.4) |
| **BMI (kg/m<sup>2</sup>)** | 27.0 (6.0) | 28.0 (6.8) | 27.9 (6.8) |
| <18.5 | 27477 (40.8) | 1792 (36.3) | 502 (36.8) |
| 18.5-24.9 | 1372 (2.0) | 121 (2.4) | 36 (2.6) |
| 25-29.9 | 22390 (33.2) | 1493 (30.2) | 405 (29.7) |

|  | United States |  |  |
| --- | --- | --- | --- |
|  | Willing<br>(*n*=67344) | Unsure<br>(*n*=4943) | Unwilling<br>(*n*=1363) |
| ≥30 | 16105 (23.9) | 1537 (31.1) | 420 (30.8) |
| **Comorbidities** |  |  |  |
| Diabetes | 3626 (5.4) | 240 (4.9) | 76 (5.6) |
| Heart Disease | 5296 (7.9) | 318 (6.4) | 92 (6.7) |
| Lung Disease | 1907 (2.8) | 168 (3.4) | 43 (3.2) |
| Kidney Disease | 1290 (1.9) | 97 (2.0) | 34 (2.5) |
| Cancer | 1955 (2.9) | 113 (2.3) | 19 (1.4) |
| **Education** | 45.9 (18.4) | 37.5 (17.5) | 34.4 (16.3) |
| **Income** | 81764 (31778) | 73783 (28354) | 70161 (25705) |
| **Current/prior smoking** | 19582 (29.1) | 1542 (31.2) | 411 (30.2) |
| **Healthcare worker** | 1833 (2.7) | 301 (6.1) | 153 (11.2) |
| **Prior COVID-19** | 3734 (5.5) | 702 (14.2) | 263 (19.3) |

|  | United Kingdom |  |  |
| --- | --- | --- | --- |
|  | Willing<br>(*n*=1093110) | Unsure<br>(*n*=52021) | Unwilling<br>(*n*=9857) |
| **Age (years)** | 54.9 (15.0) | 46.1 (14.7) | 45.0 (15.3) |
| <25 | 42646 (3.9) | 4609 (8.9) | 855 (8.7) |
| 25-34 | 81363 (7.4) | 7383 (14.2) | 1907 (19.3) |
| 35-44 | 142917 (13.1) | 11521 (22.1) | 2320 (23.5) |
| 45-54 | 218009 (19.9) | 12823 (24.6) | 1979 (20.1) |
| 55-64 | 285250 (26.1) | 10027 (19.3) | 1702 (17.3) |
| ≥65 | 322925 (29.5) | 5658 (10.9) | 1094 (11.1) |
| **Female sex** | 631477 (57.8) | 35134 (67.5) | 6467 (65.6) |
| **Race/ethnicity** |  |  |  |
| White | 1053810 (96.4) | 47663 (91.6) | 9071 (92.0) |
| Black | 7171 (0.7) | 1340 (2.6) | 276 (2.8) |
| South Asian | 13712 (1.3) | 1258 (2.4) | 229 (2.3) |
| Middle East/East Asian | 6175 (0.6) | 677 (1.3) | 94 (1.0) |
| More than one/other | 12242 (1.1) | 1083 (2.1) | 187 (1.9) |
| **BMI (kg/m<sup>2</sup>)** | 26.6 (6.0) | 26.4 (6.4) | 26.1 (6.4) |
| <18.5 | 470397 (43.0) | 23423 (45.0) | 4452 (45.2) |
| 18.5-24.9 | 28162 (2.6) | 2122 (4.1) | 478 (4.8) |
| 25-29.9 | 368595 (33.7) | 15604 (30.0) | 2972 (30.2) |
| ≥30 | 225956 (20.7) | 10872 (20.9) | 1955 (19.8) |
|  | Willing<br>(n=1093110) | Unsure<br>(n=52021) | Unwilling<br>(n=9857) |
| **Comorbidities** |  |  |  |
| Diabetes | 36909 (3.4) | 1004 (1.9) | 198 (2.0) |
| Heart Disease | 42258 (3.9) | 979 (1.9) | 191 (1.9) |
| Lung Disease | 22022 (2.0) | 672 (1.3) | 127 (1.3) |
| Kidney Disease | 9459 (0.9) | 315 (0.6) | 61 (0.6) |
| Cancer | 14540 (1.9) | 311 (0.9) | 88 (1.3) |
| **Index of multiple deprivation, education score** | 7.1 (2.5) | 6.5 (2.7) | 6.4 (2.7) |
| **Index of multiple deprivation, income score** | 7.0 (2.5) | 6.4 (2.7) | 6.3 (2.7) |
| **Current/prior smoking** | 242643 (22.2) | 12671 (24.4) | 2404 (24.4) |
| **Healthcare worker** | 24300 (2.2) | 1972 (3.8) | 653 (6.6) |
| **Prior COVID-19** | 62681 (5.7) | 4583 (8.8) | 1034 (10.5) |

All racial categories were defined as each respective race not of Hispanic or Latino ancestry. Census-level data on education used the education score and income score for the Index of Multiple Deprivation, respectively. N (percentages) presented for categorical variables. Values are means (SD) for continuous variables. Values of polytomous variables may not sum to 100% due to rounding. Abbreviations: BMI (body mass index), m (meter), kg (kilogram)

**Table S4.** Vaccine hesitancy by geographical region according to country of enrollment.

|  | United States |  |  |  |
| --- | --- | --- | --- | --- |
|  | Northeast | Midwest | South | West |
| **Number of individuals hesitant or unsure / total** | 1174/16365 | 1133/13093 | 1931/17633 | 1969/24474 |
| **Age-adjusted OR (95% CI)<sup>1</sup>** | 1.0 (ref.) | 1.16 (1.06 to 1.26) | 1.43 (1.33 to 1.54) | 1.16 (1.08 to 1.25) |
| **Multivariable-adjusted OR (95% CI)<sup>2</sup>** | 1.0 (ref.) | 1.12 (1.03 to 1.23) | 1.31 (1.21 to 1.41) | 1.13 (1.05 to 1.23) |
| **Multivariable-adjusted OR (95% CI)<sup>3</sup>** | 1.0 (ref.) | 1.03 (0.94 to 1.12) | 1.24 (1.14 to 1.34) | 1.04 (0.96 to 1.12) |

<sup>1</sup>Conditioned upon age and date of study entry
<sup>2</sup>Additional conditioning upon sex and adjustment for race/ethnicity, personal history of diabetes, heart disease, lung disease, kidney disease, current smoking status, body mass index, and prior reported history of COVID-19 infection
<sup>3</sup>Additional adjustment for frontline healthcare worker status and education and income at the community level

|  | United Kingdom |  |  |  |
| --- | --- | --- | --- | --- |
|  | England | Northern Ireland | Scotland | Wales |
| **Number of individuals hesitant or unsure / total** | 50580/952267 | 434/5994 | 2744/56232 | 2581/49811 |
| **Age-adjusted OR (95% CI)<sup>1</sup>** | 1.0 (ref.) | 1.38 (1.25 to 1.51) | 0.96 (0.93 to 1.00) | 1.10 (1.06 to 1.15) |
| **Multivariable-adjusted OR (95% CI)<sup>2</sup>** | 1.0 (ref.) | 1.44 (1.31 to 1.58) | 1.00 (0.96 to 1.04) | 1.13 (1.08 to 1.18) |
| **Multivariable-adjusted OR (95% CI)<sup>3</sup>** | 1.0 (ref.) | 1.42 (1.30 to 1.57) | 1.00 (0.96 to 1.04) | 1.11 (1.07 to 1.16) |

<sup>1</sup>Conditioned upon age and date of study entry
<sup>2</sup>Additional conditioning upon sex and adjustment for race/ethnicity, personal history of diabetes, heart disease, lung disease, kidney disease, current smoking status, body mass index, and prior reported history of COVID-19 infection
<sup>3</sup>Additional adjustment for frontline healthcare worker status and education and income at the community level
Abbreviations: CI (confidence interval), OR (odds ratio)

**Table S5.** Reasons for vaccine hesitancy by race and ethnicity according to country of enrollment.

| Reason Cited (%) | United States |  |  |  |  |
| --- | --- | --- | --- | --- | --- |
|  | White<br>(n=4715) | Black<br>(n=611) | Hispanic<br>(n=505) | Asian<br>(n=309) | More than<br>one/other<br>(n=166) |
| Concerned about the long-term side effects | 51.8 | 57.3 | 54.9 | 49.5 | 47.0 |
| Concerned about adverse reaction | 44.7 | 54.2 | 51.9 | 47.2 | 45.8 |
| Do not know enough about it | 41.7 | 50.6 | 50.9 | 42.4 | 41.6 |
| Personal belief/philosophical reasons | 8.8 | 14.1 | 13.1 | 7.8 | 10.8 |
| Illness/Medication | 8.3 | 8.7 | 7.5 | 7.1 | 12.1 |
| Do not think it is necessary | 6.7 | 2.5 | 4.2 | 3.9 | 10.8 |
| Do not think it will be available to me | 5.0 | 4.6 | 6.5 | 7.1 | 6.0 |
| Prefer not to say | 4.3 | 6.2 | 4.2 | 4.2 | 4.2 |
| Do not think it will work | 3.9 | 3.8 | 4.6 | 2.9 | 7.2 |
| Pregnancy/breastfeeding | 2.4 | 0.7 | 2.8 | 4.5 | 1.2 |
| Religious reasons | 1.9 | 1.3 | 3.6 | 2.6 | 2.4 |
| I took part in a vaccine trial | 0.3 | 0.3 | 0.4 | 0.3 | 0.6 |
| Other | 9.8 | 9.8 | 6.1 | 9.4 | 13.9 |

| Reason Cited (%) | United Kingdom |  |  |  |  |
| --- | --- | --- | --- | --- | --- |
|  | White<br>(n=56775) | Black<br>(n=1617) | South Asian<br>(n=1487) | Middle<br>East/East<br>Asian<br>(n=771) | More than<br>one/other<br>(n=1270) |
| Concerned about the long-term side effects | 49.9 | 52.9 | 50.9 | 55.6 | 51.3 |
| Concerned about adverse reaction | 30.2 | 39.3 | 36.5 | 37.2 | 34.4 |
| Do not know enough about it | 36.7 | 45.0 | 41.9 | 39.8 | 36.0 |
| Personal belief/philosophical reasons | 5.4 | 7.3 | 6.3 | 4.0 | 5.0 |
| Illness/Medication | 5.8 | 5.9 | 5.5 | 5.3 | 4.9 |
| Do not think it is necessary | 7.5 | 5.6 | 5.9 | 5.1 | 7.6 |
| Do not think it will be available to me | 9.0 | 7.2 | 9.8 | 8.4 | 10.9 |
| Prefer not to say | 5.8 | 8.0 | 8.3 | 6.2 | 5.5 |
| Do not think it will work | 3.0 | 2.8 | 4.7 | 3.4 | 3.7 |
| Pregnancy/breastfeeding | 7.6 | 4.8 | 5.9 | 6.0 | 6.3 |
| Religious reasons | 0.5 | 1.9 | 1.8 | 1.0 | 1.0 |
| I took part in a vaccine trial | 0.4 | 0.1 | 0.4 | 0.4 | 0.3 |
| Other | 8.2 | 6.2 | 5.1 | 7.4 | 9.0 |

Proportions are calculated within racial and ethnic categories among individuals expressing vaccine unwillingness or unsure if they will obtain one. Participants were allowed to check more than one reason.

**Table S6.**
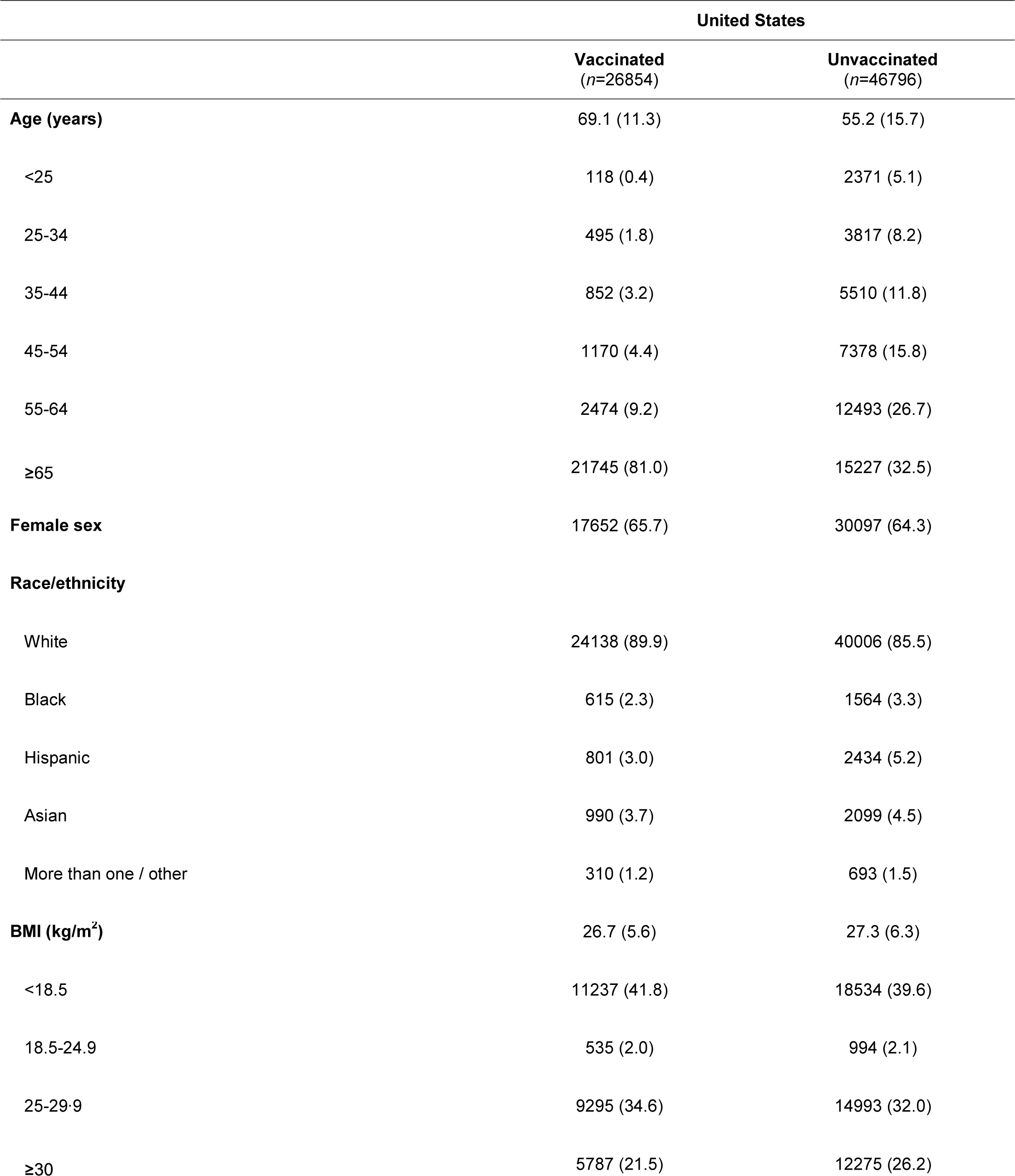
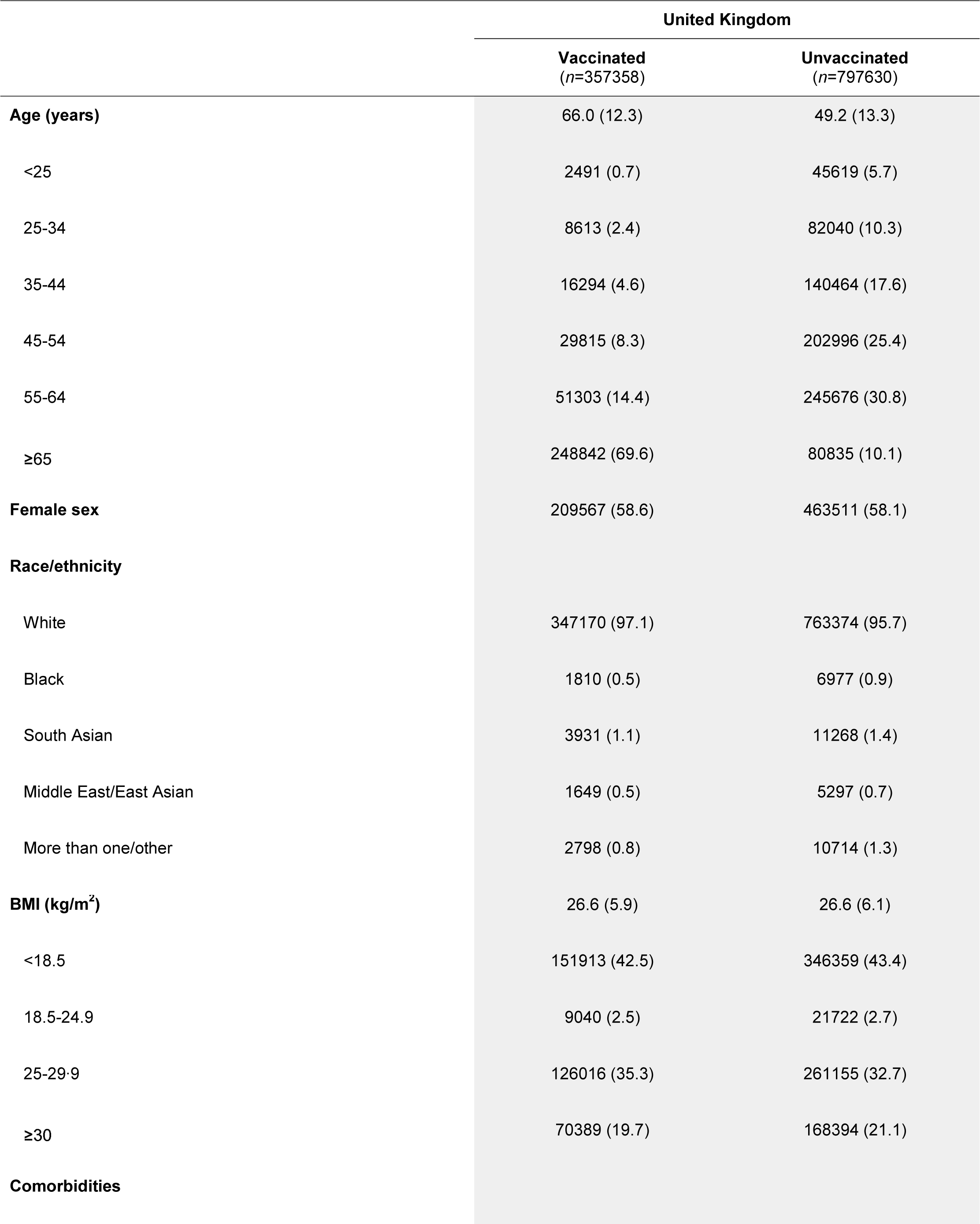
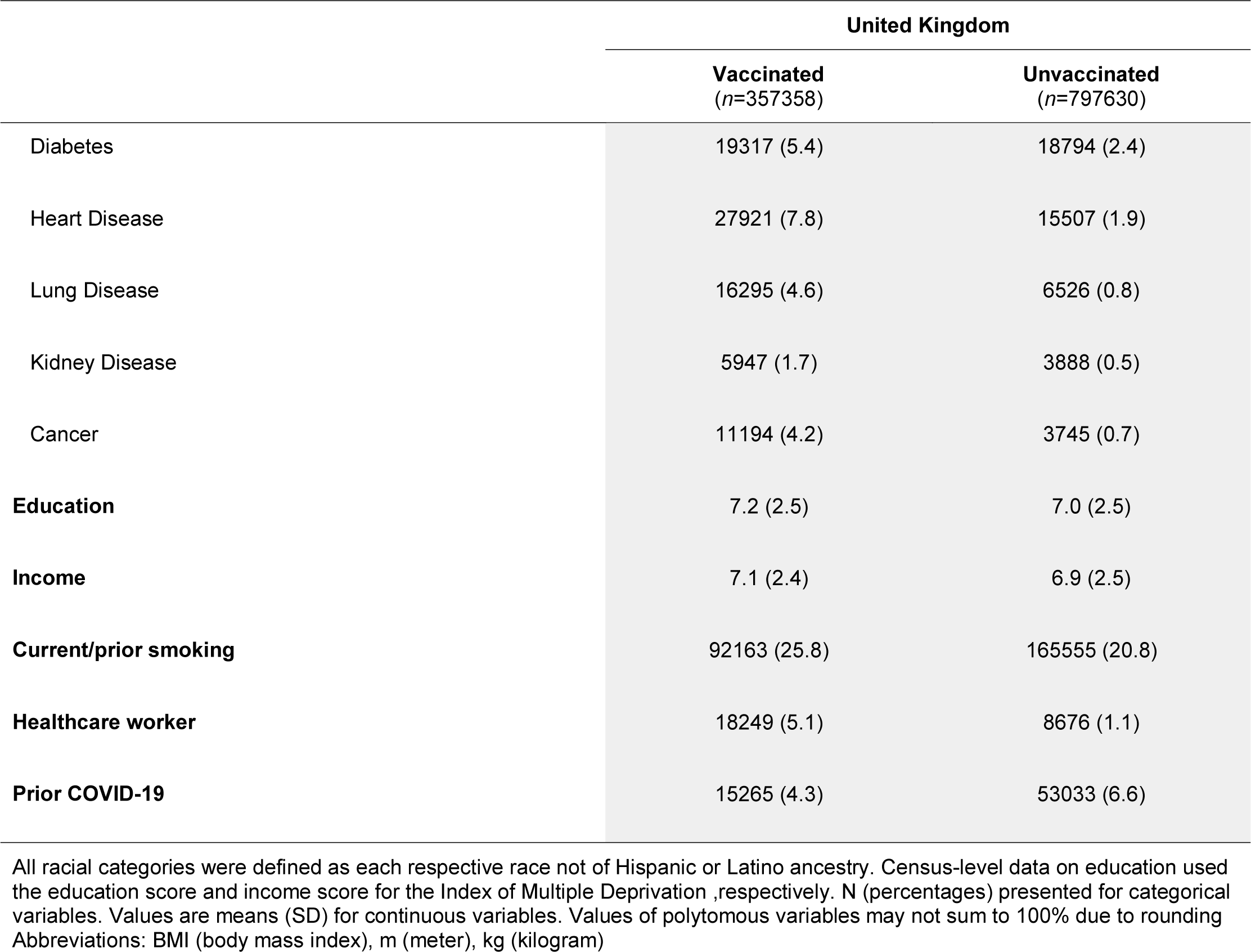
Baseline characteristics of study participants by vaccination status and country of enrollment.

**Table S7.** Vaccine uptake by geographical region according to country of enrollment.

|  | United States |  |  |  |
| --- | --- | --- | --- | --- |
|  | Northeast | Midwest | South | West |
| **Number of individuals receiving a vaccine / total** | 2942/16365 | 2569/13093 | 4541/17633 | 6748/24474 |
| **Age-adjusted OR (95% CI)<sup>1</sup>** | 1.0 (ref.) | 1.13 (1.07 to 1.19) | 1.69 (1.61 to 1.77) | 1.52 (1.46 to 1.59) |
| **Multivariable-adjusted OR (95% CI)<sup>2</sup>** | 1.0 (ref.) | 1.14 (1.08 to 1.21) | 1.70 (1.62 to 1.79) | 1.52 (1.45 to 1.59) |
| **Multivariable-adjusted OR (95% CI)<sup>3</sup>** | 1.0 (ref.) | 1.14 (1.08 to 1.21) | 1.73 (1.65 to 1.82) | 1.55 (1.48 to 1.62) |

<sup>1</sup>Conditioned upon age and date of study entry
<sup>2</sup>Additional conditioning upon sex and adjustment for race/ethnicity, personal history of diabetes, heart disease, lung disease, kidney disease, current smoking status, body mass index, and prior reported history of COVID-19 infection
<sup>3</sup>Additional adjustment for frontline healthcare worker status and education and income at the community level

|  | United Kingdom |  |  |  |
| --- | --- | --- | --- | --- |
|  | England | Northern Ireland | Scotland | Wales |
| **Number of individuals receiving a vaccine / total** | 151200/952267 | 884/5994 | 4290/56232 | 6770/49811 |
| **Age-adjusted OR (95% CI)<sup>1</sup>** | 1.0 (ref.) | 0.92 (0.86 to 0.98) | 0.42 (0.40 to 0.43) | 0.72 (0.70 to 0.74) |
| **Multivariable-adjusted OR (95% CI)<sup>2</sup>** | 1.0 (ref.) | 0.91 (0.85 to 0.97) | 0.42 (0.40 to 0.42) | 0.72 (0.70 to 0.73) |
| **Multivariable-adjusted OR (95% CI)<sup>3</sup>** | 1.0 (ref.) | 0.89 (0.83 to 0.95) | 0.41 (0.40 to 0.42) | 0.71 (0.70 to 0.73) |

<sup>1</sup>Conditioned upon age and date of study entry
<sup>2</sup>Additional conditioning upon sex and adjustment for race/ethnicity, personal history of diabetes, heart disease, lung disease, kidney disease, current smoking status, body mass index, and prior reported history of COVID-19 infection
<sup>3</sup>Additional adjustment for frontline healthcare worker status and education and income at the community level
Abbreviations: CI (confidence interval), OR (odds ratio). Data shown through February 1, 2021.

**Table S8.** Localized symptoms among vaccinated participants according to country of enrollment.

|  | United States |  |  |  |  |
| --- | --- | --- | --- | --- | --- |
|  | White<br>(n=32316) | Black<br>(n=811) | Hispanic<br>(n=1198) | Asian<br>(n=1328) | More than<br>one/other<br>(n=431) |
| Pain | 9115 (28.2) | 228 (28.1) | 400 (33.4) | 492 (37.0) | 134 (31.1) |
| Redness | 708 (2.2) | 19 (2.3) | 36 (3.0) | 30 (2.3) | 12 (2.8) |
| Swelling | 1427 (4.4) | 73 (9.0) | 78 (6.5) | 98 (7.4) | 27 (6.3) |
| Swollen lymph nodes | 159 (0.5) | 11 (1.4) | 13 (1.1) | 6 (0.5) | 4 (0.9) |
| Warmth | 1463 (4.5) | 46 (5.7) | 94 (7.8) | 76 (5.7) | 38 (8.8) |
| Bruising | 78 (1.1) | 1 (0.5) | 10 (4.6) | 3 (1.4) | 2 (2.1) |
| Itchiness | 457 (1.4) | 27 (3.3) | 22 (1.8) | 29 (2.2) | 4 (0.9) |
| Tenderness | 11950 (37.0) | 281 (34.6) | 466 (38.9) | 528 (39.8) | 157 (36.4) |
|  | United Kingdom |  |  |  |  |
|  | White<br>(n=393456) | Black<br>(n=2218) | South Asian<br>(n=5092) | Middle East/East<br>Asian<br>(n=2051) | More than<br>one/other<br>(n=3378) |
| Pain | 53666 (13.6) | 594 (26.8) | 1662 (32.6) | 514 (25.1) | 800 (23.7) |
| Redness | 6387 (1.6) | 56 (2.5) | 131 (2.6) | 45 (2.2) | 82 (2.4) |
| Swelling | 10144 (2.6) | 158 (7.1) | 302 (5.9) | 81 (3.9) | 160 (4.7) |
| Swollen lymph nodes | 1558 (0.4) | 23 (1.0) | 46 (0.9) | 11 (0.5) | 22 (0.7) |
| Warmth | 15787 (4.0) | 164 (7.4) | 300 (5.9) | 119 (5.8) | 171 (5.1) |
| Bruising | 2095 (1.7) | 12 (2.0) | 18 (1.5) | 12 (2.2) | 25 (2.6) |
| Itchiness | 4029 (1.0) | 38 (1.7) | 78 (1.5) | 29 (1.4) | 58 (1.7) |
| Tenderness | 111833 (28.4) | 760 (34.3) | 1559 (30.6) | 592 (28.9) | 1101 (32.6) |

Proportions are calculated within racial and ethnic categories among individuals expressing vaccine unwillingness or unsure if they will obtain one. Participants were allowed to check more than one reason.

## Notes

### Author Declarations

This research study was approved by the Mass General Brigham Human Research Committee (Institutional Review Board Protocol 2020P000909) and King's College London Ethics Committee (REMAS ID 18210).

